# Short-Term Safety of Low-Dose Creatine Hydrochloride: A 28-Day Single-Arm Pilot Study

**DOI:** 10.64898/2026.04.10.26349886

**Authors:** Jon C Wagner, Sergej Ostojic, William Faulkner, Mark Faulkner

**Affiliations:** Third Quarter Consulting 13004 Binney Street, Omaha, NE 68164, USA; Texas Tech University, 2500 Broadway, Lubbock, TX 79409; University of Agder Universitetsveien 25, 4630 Kristiansand, Norway; Vireo Systems, Madison, TN USA 810 Royal Parkway, Nashville, TN 37214 USA

**Keywords:** Creatine hydrochloride, supplementation, safety, clinical biomarkers, pilot study

## Abstract

**Background:** Creatine monohydrate (typically 5-20 g/day) has a well-established safety profile across diverse populations. Creatine hydrochloride (CR-HCl) is a highly soluble creatine formulation that may allow effective supplementation at substantially lower doses (750 mg - 3 g/day); however, controlled human safety data specific to CR-HCl remain limited.

**Objective:** To evaluate the short-term laboratory safety and tolerability of low-dose CR-HCl supplementation administered for 28 days in healthy adults.

**Methods:** This single-center, single-arm, single-blind pilot safety study enrolled 11 healthy adults (10 females, 1 male; mean age 44.6 ± 7.2 years). Participants consumed 750 mg/day CR-HCl for 28 consecutive days while maintaining their usual diet and physical activity patterns.

Fasting blood and urine samples were collected at baseline and Day 28. Laboratory assessments included hematological, lipid, and clinical chemistry biomarkers. Pre–post changes were evaluated using paired parametric and nonparametric tests, baseline-adjusted regression models, bootstrap confidence intervals, and false discovery rate (FDR) correction.

**Results:** All participants completed the intervention. No clinically meaningful changes were observed in lipid parameters, hematologic indices, renal markers, or most chemistry analytes after adjustment for multiple comparisons. Fasting glucose increased modestly (8.1 mg/dL) prior to multiplicity adjustment but was not statistically significant after FDR correction and remained within reference ranges. Serum bicarbonate decreased slightly (2.4 mmol/L); although statistically detectable in parametric analysis, values remained within physiological limits and were not consistently supported by nonparametric testing.

**Conclusions:** Supplementation with 750 mg/day CR-HCl for 28 days was well tolerated and was not associated with clinically meaningful alterations in routine laboratory biomarkers. These preliminary findings support the short-term tolerability of low-dose CR-HCl and provide a basis for larger randomized, placebo-controlled studies to further evaluate its safety profile.

## Introduction

Creatine supplementation is one of the most extensively studied nutritional strategies for enhancing physical performance, supporting recovery, and promoting aspects of metabolic and neuromuscular health. Traditionally, most clinical research has focused on creatine monohydrate, typically administered at doses ranging from 5 to 20 g/day depending on the loading or maintenance protocol. Across numerous randomized trials and long-term observational studies, creatine monohydrate has demonstrated a strong safety profile in healthy individuals as well as in clinical populations, with no consistent evidence of adverse effects on renal, hepatic, cardiovascular, or hematological function when used within recommended ranges.^1-4^ In recent years, alternative creatine formulations have been developed with the aim of improving physicochemical properties such as solubility, gastrointestinal tolerability, and dosing efficiency. Creatine hydrochloride (CR-HCl) represents one such formulation. Due to its markedly higher aqueous solubility compared with creatine monohydrate, CR-HCl may allow effective supplementation at substantially lower doses, typically in the range of 750 mg to 3 g/day.^5-8^ Improved solubility may also facilitate easier formulation in liquid preparations and reduce the likelihood of gastrointestinal discomfort sometimes reported with higher-dose creatine monohydrate protocols. Despite the growing commercial availability and use of CR-HCl, comparatively few controlled studies have systematically evaluated its clinical safety profile. While anecdotal reports and preliminary investigations suggest good tolerability, there remains limited laboratory-based evidence examining the short-term effects of low-dose CR-HCl supplementation on routine biochemical, hematological, and metabolic markers in humans.

Given the increasing adoption of lower-dose creatine strategies and the emergence of alternative formulations, establishing formulation-specific safety data is important for both clinical practice and product development. Therefore, the purpose of the present pilot study was to evaluate the short-term laboratory safety of creatine hydrochloride supplementation administered at a daily dose of 750 mg for 28 days in healthy adults. A comprehensive panel of hematological, lipid, and clinical chemistry biomarkers was assessed to determine whether low-dose CR-HCl supplementation induces measurable changes in physiological safety indicators over the intervention period.

## Methods

This single-center, single-arm, single-blind pilot study was conducted at Princeton Consumer Research Corp. (Raritan, NJ, USA) between May 30 and July 3, 2024. The study protocol was reviewed and approved by the Univo Institutional Review Board (IRB2024-SP029) on May 2, 2024, and all participants provided written informed consent prior to enrollment. The study was funded by Vireo Systems (Nashville, TN, USA). Healthy adults aged 18–60 years were eligible to participate if they agreed to maintain their usual diet and physical activity patterns throughout the study period and to fast for at least 12 hours prior to scheduled blood collections.

Participants were excluded if they had consumed any creatine-containing supplements within the previous 30 days or had a history of clinically significant medical conditions that could affect study outcomes, including diabetes mellitus, hypertension requiring treatment, renal disease, hepatitis, HIV/AIDS, or malignancy within the past five years. Twelve individuals were screened for eligibility, and eleven participants were enrolled in the study. No formal power calculation was performed, as the study was designed as an exploratory pilot trial intended to generate preliminary safety data. The sample size was therefore determined based on feasibility considerations. Participants were required to consume 750 mg of CR-HCl powder (CONCRET®, Vireo Systems, Nashville, TN, USA) once daily for 28 consecutive days. The dose of 750 mg of CR-HCl was chosen for this trial because it is the commercially recommended dose. The supplement was taken after the first meal of the day. Participants were blinded to the specific identity of the test product. Compliance and Adverse Events were monitored via diary logs kept by study participants. Unused supplement on Visit 3 (Day 28) was also used to determine compliance. Morning fasting blood and urine samples were collected at baseline (Day 0) and after 28 days of supplementation (Day 28). Laboratory assessments included a complete blood count (CBC) with differential, lipid panel, glucose, renal function markers (blood urea nitrogen, serum creatinine, estimated glomerular filtration rate, and BUN/creatinine ratio), electrolytes (sodium, potassium, chloride, bicarbonate, calcium), protein status (total protein, albumin, globulin), bilirubin, and hepatic enzymes (alkaline phosphatase, AST, ALT). Statistical analyses were performed to evaluate changes between baseline and Day 28. Paired t-tests and Wilcoxon signed-rank tests were used to assess pre–post differences. Baseline-adjusted changes were additionally evaluated using ANCOVA-style regression models. Paired bootstrap resampling (20,000 iterations) was applied to generate 95% confidence intervals (CIs) for mean changes. To account for multiple comparisons across laboratory outcomes, false discovery rate (FDR) correction was applied. Given the exploratory nature of the study and the small sample size, emphasis was placed on effect magnitude, confidence intervals, and clinical reference ranges rather than hypothesis testing alone. The risk of analytical bias was minimal, as outcomes were based on standardized laboratory measurements and objective statistical procedures

## Results

All eleven enrolled participants completed the 28-day supplementation period. The cohort was predominantly female (90.9%), with a mean age of 44.6 ± 7.2 years. No participants withdrew due to adverse events, and no clinically significant safety concerns were observed during the study. Biochemical biomarkers across the intervention period are summarized in Table 1 (Blood chemistry analysis) and Table 2 (CDC and lipid profile).

**Table 1:**
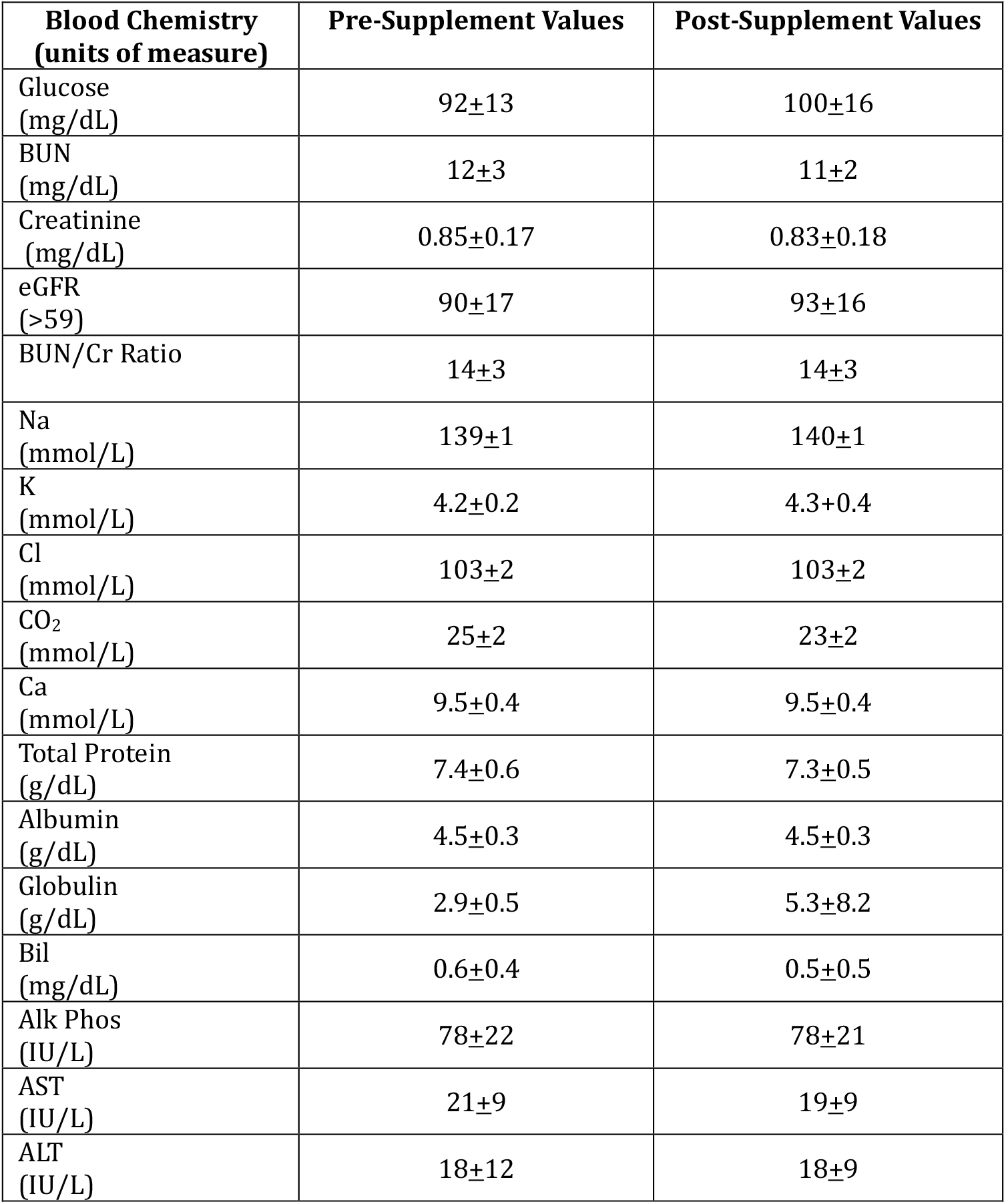
Blood Chemistry Values Prior to and at the Conclusion of Creatine HCl Supplementation.

**Table 2:**
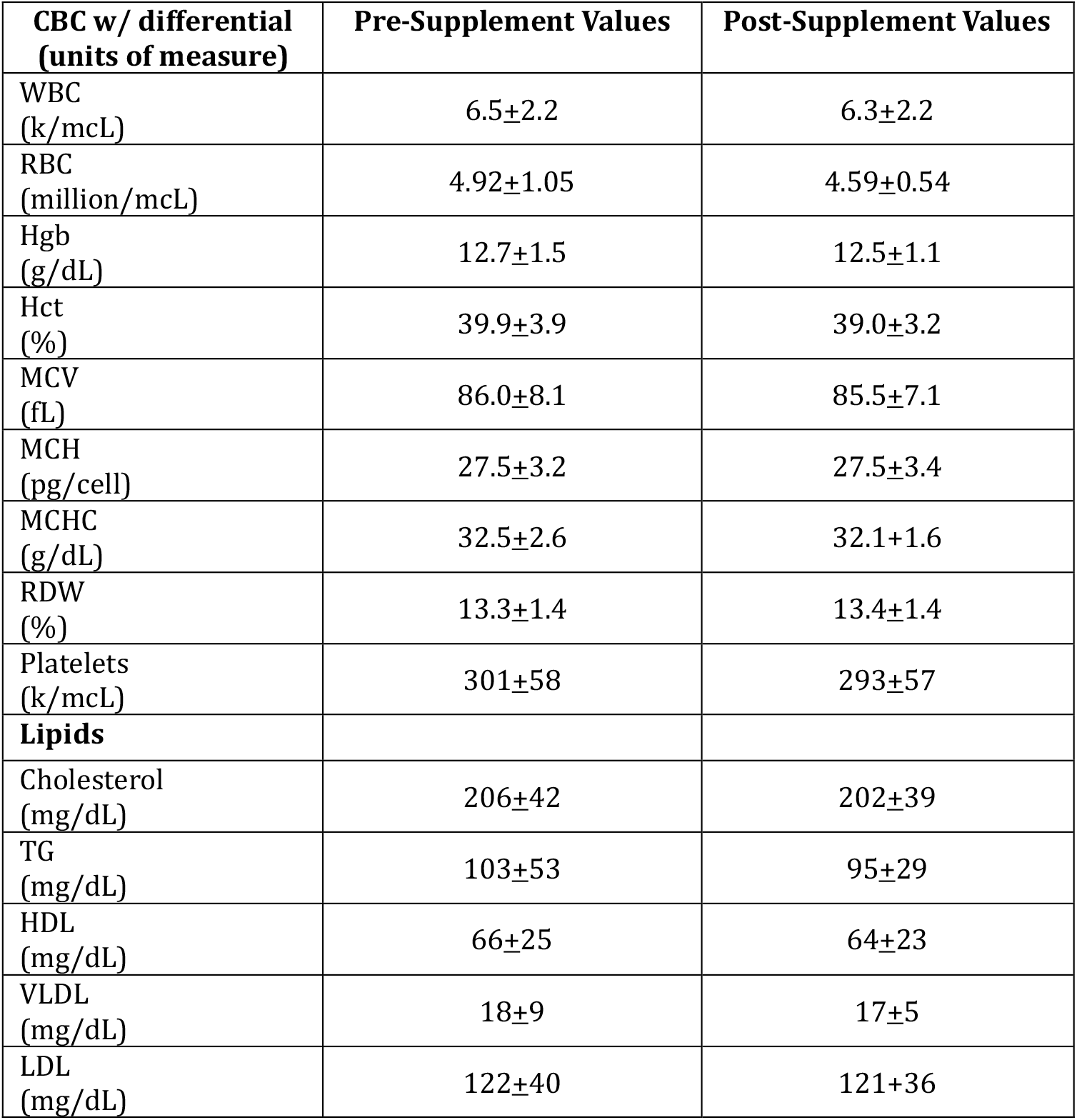
CBC and Lipid Profile Pre and Post Creatine HCl Supplementation.

Blood urea nitrogen (BUN); estimated glomerular filtration rate (eGFR); Blood urea nitrogen – creatinine ratio (BUN/Cr); sodium (Na); potassium (K); chloride (Cl); carbon dioxide (CO_2_); calcium (Ca); bilirubin (Bil); alkaline phosphatase (Alk Phos); aspartate aminotransferase (AST); alanine aminotransferase (ALT) Complete blood count (CBC); white blood cell (WBC); red blood cell (RBC); hemoglobin (HgB); hematocrit (Hct); mean corpuscular volume (MCV); mean corpuscular hemoglobin (MCH); mean corpuscular hemoglobin concentration (MCHC); red cell distribution width (RDW); triglycerides (TG); high-density lipoprotein (HDL); very low-density lipoprotein (VLDL); low-density lipoprotein (LDL) Mean changes in lipid parameters were small in magnitude. Total cholesterol, triglycerides, HDL-C, LDL-C, and VLDL-C decreased by 4.3 mg/dL, 7.7 mg/dL, 2.1 mg/dL, 1.1 mg/dL, and 1.1 mg/dL, respectively. None of these changes reached statistical significance after FDR correction. Effect sizes ranged from trivial to small, and bootstrap 95% CIs for all lipid outcomes included zero, indicating no detectable systematic shift. ANCOVA-style models demonstrated strong within-subject stability (R^2^ = 0.82-0.98), suggesting that 28-day lipid values were largely predicted by baseline levels rather than influenced by supplementation. Importantly, no participants developed clinically abnormal lipid values during the study. Complete blood count indices remained stable throughout the intervention. No statistically significant pre-post changes were observed in leukocyte count, erythrocyte indices, or platelet count after adjustment for multiple comparisons. Effect sizes were uniformly small, and bootstrap confidence intervals encompassed zero for all variables. No participant demonstrated laboratory evidence of hemoconcentration, cytopenia, or an inflammatory response. Across the 17 chemistry analytes evaluated, most exhibited minimal mean change from baseline. Fasting glucose increased modestly (8.1 mg/dL) before multiplicity adjustment but did not remain statistically significant after FDR correction. All values remained within standard reference ranges, and no participant met criteria for impaired fasting glucose.

Serum bicarbonate decreased by 2.4 mmol/L; this change remained statistically significant in parametric testing after FDR correction but was not consistently supported by nonparametric analysis. Importantly, all post-supplementation values remained within physiological limits, and no participant demonstrated laboratory evidence of metabolic acidosis. Renal markers, including serum creatinine and 24-hour urinary creatinine, remained stable with no clinically meaningful changes. Serum creatine kinase, amylase, and lipase also showed no significant differences from baseline. Collectively, the laboratory findings were consistent with expected intra-individual biological variability over the 28-day study period.

## Discussion

This pilot study evaluated the short-term laboratory safety of low-dose CR-HCl supplementation in healthy adults. Across multiple physiological domains, including lipid metabolism, hematologic indices, renal function, and clinical chemistry, no clinically meaningful adverse changes were observed. Laboratory values remained within reference ranges, and the overall pattern of findings was consistent with expected intra-individual biological variability. Given the absence of a comparator group and the small sample size, the findings should be interpreted as preliminary tolerability observations rather than definitive safety evidence. These observations are broadly consistent with the extensive literature examining the safety of creatine supplementation. Most available data derive from studies using creatine monohydrate, which has been evaluated across a wide range of dosing strategies and populations. Randomized trials and long-term observational studies have repeatedly shown that creatine supplementation, even at substantially higher doses than used in the present study, does not produce clinically meaningful alterations in renal, hepatic, hematologic, or metabolic biomarkers in healthy individuals.^1,3,4^ Systematic reviews and meta-analyses similarly conclude that creatine supplementation is generally well tolerated when consumed within recommended ranges.^4,6^ Although CR-HCl differs chemically from creatine monohydrate and is typically administered at lower doses due to its higher aqueous solubility, the present findings suggest that its short-term physiological safety profile is comparable.^5-7^ Particular attention is often directed toward renal safety when evaluating creatine supplementation. Concerns have historically arisen from the role of creatine metabolism in generating creatinine, a commonly used biomarker of kidney function. However, controlled studies have consistently demonstrated that creatine supplementation does not impair renal function in healthy individuals.^9,10^ The current findings are in line with this body of evidence and provide additional reassurance that low-dose CR-HCl does not adversely affect routine renal laboratory markers over short durations. A small reduction in serum bicarbonate was observed in parametric analysis, although the magnitude of change was modest and remained well within physiological reference ranges. Moreover, the effect was not consistently supported by nonparametric testing and was not accompanied by other laboratory indicators of acid-base disturbance. Variability of this magnitude is commonly observed in repeated biochemical measurements and likely reflects normal physiological fluctuation rather than a treatment-related effect.

Additional context can be drawn from post-marketing safety surveillance data. The U.S. Food and Drug Administration’s Center for Food Safety and Applied Nutrition Adverse Event Reporting System (CAERS) collects voluntary reports of adverse events associated with foods and dietary supplements.^11^ A search of the CAERS database identified 11 reports related specifically to CR-HCl among 229,467 total entries recorded between 1989 and 2025 (REF), corresponding to an estimated prevalence of approximately 0.0048% or roughly 0.41 reports per year. By comparison, all forms of creatine supplementation accounted for 207 reports during the same time period, representing approximately 7.67 reports per year. Although CAERS data cannot establish causality and often lack detailed clinical context, the low frequency of reported adverse events provides supportive real-world evidence consistent with the laboratory safety findings observed in the present study.

Several strengths should be considered when interpreting these results. The study incorporated a comprehensive panel of laboratory biomarkers spanning multiple physiological systems, standardized testing procedures performed at a single accredited laboratory, and multiple complementary statistical approaches including parametric and nonparametric analyses, bootstrap confidence intervals, and correction for multiple comparisons. These features strengthen the reliability of the safety assessment within the constraints of an exploratory pilot design. Nevertheless, several limitations warrant consideration. The small sample size limits statistical power and the ability to detect rare or subtle physiological effects. The absence of a placebo control group precludes definitive causal attribution of observed changes. In addition, the relatively short intervention period restricts conclusions to short-term safety, and the predominantly female composition of the study cohort may limit generalizability to broader populations. Another limitation is that direct biomarkers of creatine exposure, such as plasma creatine concentrations, urinary creatine excretion, or tissue creatine levels, were not measured, preventing confirmation of systemic absorption or retention of the supplemented creatine hydrochloride. Future randomized controlled trials with larger and more diverse samples, longer intervention durations, and comparative formulations will be valuable for further characterizing the safety profile of creatine hydrochloride supplementation.

## Conclusion

In healthy adults, supplementation with 750 mg/day of creatine hydrochloride for 28 days was well tolerated and was not associated with clinically meaningful changes in routine laboratory biomarkers. These findings suggest that low-dose CR-HCl can be administered safely over the short term in healthy individuals. The results provide preliminary evidence supporting the physiological tolerability of this formulation and establish a basis for future randomized, placebo-controlled studies with larger samples and longer follow-up to further characterize its safety profile.

## Data Availability

All data produced in the present study are available upon reasonable request to the authors

## References

1. Kreider RB, Kalman DS, Antonio J, et al. International Society of Sports Nutrition position stand: safety and efficacy of creatine supplementation in exercise, sport, and medicine. J Int Soc Sports Nutr. 2017;14:18.

2. Rawson ES, Venezia AC. Use of creatine in the elderly and evidence for effects on cognitive function in young and old. Amino Acids. 2011;40(5):1349–1362.

3. Tarnopolsky MA. Creatine as a therapeutic strategy for myopathies. Amino Acids. 2011;40(5):1397–1407.

4. Antonio J, Candow DG, Forbes SC, et al. Common questions and misconceptions about creatine supplementation. J Int Soc Sports Nutr. 2021;18:13.

5. Gufford BT, Sriraghavan K, Miller NJ, et al. Physicochemical characterization of creatine salts: solubility and stability properties. AAPS PharmSciTech. 2010;11(4):1448–1454.

6. Kreider RB, Antonio J, et al. Bioavailability, efficacy, safety, and regulatory status of creatine and related compounds: a critical review. Nutrients. 2022;14:1035.

7. Vukojević V, Nikolić D, Pavlović D, et al. Solubility of creatine and its hydrochloride salts in aqueous solutions at different temperatures. J Serb Chem Soc. 2024;89:1067–1076.

8. Korovljev D, Ostojic J, Panic J, Ranisavljev M, Todorovic N, Nedeljkovic D, Kuzmanovic J, Vranes M, Stajer V, Ostojic SM. The Effects of 8-Week Creatine Hydrochloride and Creatine Ethyl Ester Supplementation on Cognition, Clinical Outcomes, and Brain Creatine Levels in Perimenopausal and Menopausal Women (CONCRET-MENOPA): A Randomized Controlled Trial. J Am Nutr Assoc. 2026 Mar-Apr;45(3):199–210. doi: 10.1080/27697061.2025.2551184. Epub 2025 Aug 25. PMID: 40854087.

9. Greenwood M, Kreider RB, Jagim AR. Creatine supplementation and kidney function: a systematic review and meta-analysis. J Int Soc Sports Nutr. 2020;17:31.

10. Naeini EK, et al. Effect of creatine supplementation on kidney function: a systematic review and meta-analysis. BMC Nephrol. 2025;26:622. doi:10.1186/s12882-025-04558-6

11. U.S. Food and Drug Administration. Center for Food Safety and Applied Nutrition Adverse Event Reporting System (CAERS). FDA; 2026.

